# Clonal Hematopoiesis in HIV and Atherosclerosis, Arterial Inflammation, and Lymph Node Metabolic Activity

**DOI:** 10.64898/2026.02.05.26345712

**Authors:** Matthew S. Durstenfeld, Katherine J. Kentoffio, Alexandra E. Teng, Shady Abohashem, Danny Li, Yifei Ma, Rebecca Hoh, Steven G. Deeks, Alexander G. Bick, Ahmed A. Tawakol, Priscilla Y. Hsue

## Abstract

**Background:** Clonal hematopoiesis of indeterminate potential (CHIP) is associated with cardiovascular disease (CVD) in the general population and is more common among people with HIV (PWH). The mechanisms by which CHIP contributes to atherosclerosis in PWH are unknown. We hypothesized that CHIP is associated with carotid atherosclerosis and arterial inflammation among PWH.

**Methods:** In a cohort study, we studied PWH ages 31-74 years with treated, suppressed HIV. CHIP mutations were detected with a validated targeted sequencing assay. Carotid intima-media thickness (IMT) was measured longitudinally with ultrasound. Aortic inflammation and lymph node activity were assessed cross-sectionally using ^18^F-FDG-PET. Inflammatory biomarkers were measured using multiplex electrochemiluminescence assay. Linear regression was employed, with adjustments for traditional and HIV-related factors.

**Results:** We included 230 PWH (52±9 years, 7% female); 32 (14%) had CHIP with median variant allele fraction of 2.8%. Common mutations were in DNM3TA (n=21) and TET2 (n=6). Age was associated with CHIP (OR 2.0 per decade older, 95% CI 1.3-3.01; p=0.002). Among 166 participants with IMT measurements (CHIP=23), CHIP was not associated with IMT (p=0.21; unchanged after adjustment). Among 80 with FDG-PET, CHIP (n=12) was not associated with arterial inflammation (p=0.89), but was associated with higher lymph node metabolic activity (p=0.017) that was attenuated in reference to background activity and adjusted for risk factors. CHIP was not associated with soluble inflammatory markers or viral persistence markers.

**Conclusions:** Among PWH, CHIP mutations were not associated with subclinical atherosclerosis, arterial inflammation, or soluble inflammatory markers but may be related to lymph node metabolic activity. The mechanism by which CHIP increases HIV-associated atherosclerosis may preferentially involve lymph nodes and merits additional evaluation.

**Visual Abstract:** 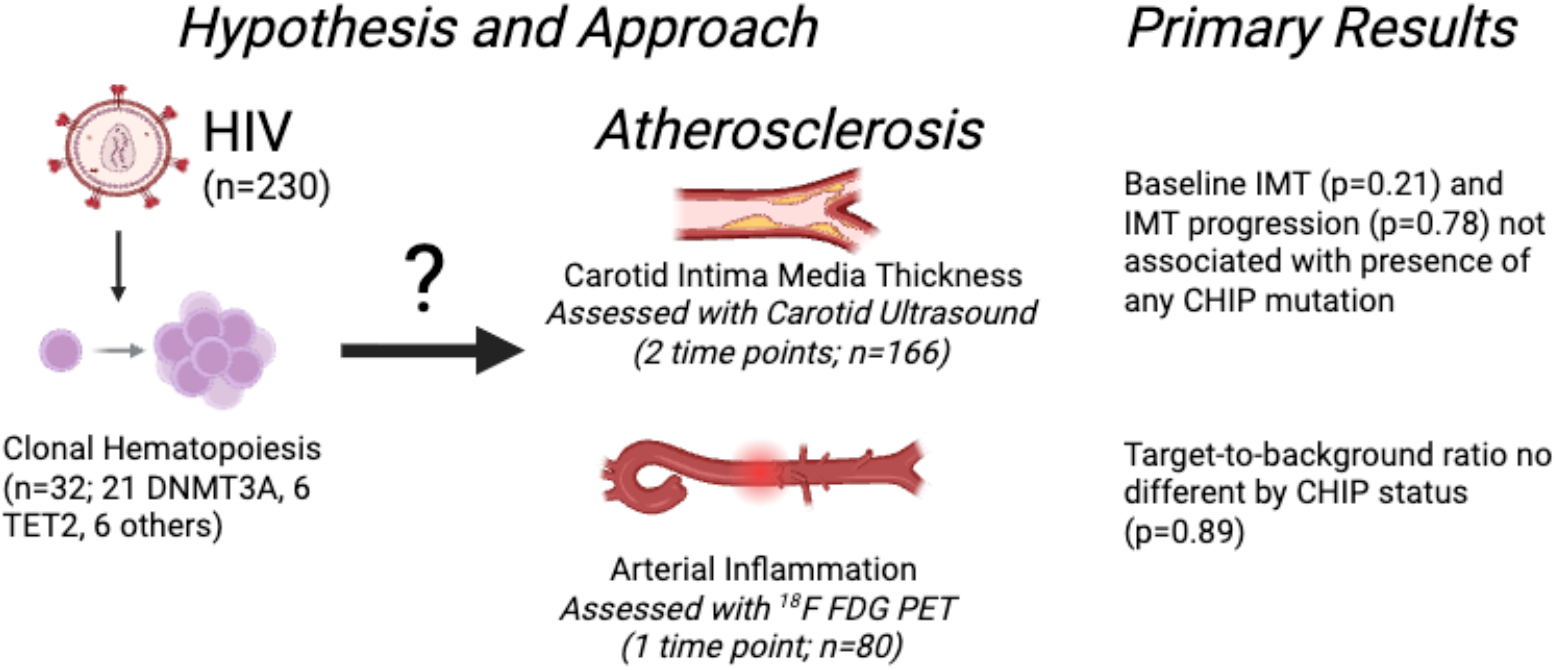

**Visual Abstract Legend:** We hypothesized that clonal hematopoeiesis of indeterminate potential (CHIP) may be a mediator of the association between HIV and atherosclerosis; we investigated this hypothesis by measuring CHIP mutations in human blood samples from people with HIV (PWH) and determining whether CHIP mutations were associated with carotid intima media thickness assessed using ultrasound (longitudinal, n=166) and aortic inflammation using ^18^F-FDG-PET-CT (cross-sectional, n=80). In our study of 230 PWH (32 with CHIP), we did not find evidence for an association between the presence of any CHIP mutation and either carotid atherosclerosis or arterial inflammation.

**Condensed Abstract:** People with HIV (PWH) ave increased risk of clonal hematopoiesis of indeterminate potential (CHIP) and atherosclerosis, but whether clonal hematopoiesis accelerates atherosclerosis among PWH is unknown. We hypothesized that CHIP is associated with carotid atherosclerosis and arterial inflammation among PWH. In a cohort study that conducted longitudinal carotid ultrasound and 18F-FDG-PET, we studied PWH ages 31-74 years with treated, suppressed HIV. We measured CHIP mutations with a validated targeted sequencing assay. We included 230 PWH (52±9 years, 7% female); 32 (14%) had CHIP (66% DNMT3A) with median variant allele fraction of 2.8%. Overall, CHIP was not associated with carotid intima media thickness or arterial inflammation assessed with FDG PET. CHIP was associated with increased lymph node metabolic activity, a hypothesis-generating finding. Among PWH, CHIP was not associated with markers of subclinical atherosclerosis.

## Introduction

HIV is associated with approximately double the risk of cardiovascular disease, including elevated risk of atherosclerotic cardiovascular disease such as myocardial infarction and stroke, as well as heart failure and arrhythmias (1,2). Multiple mechanistic pathways contribute to the increased cardiovascular risk among people with HVI (PWH), many of which result in immune dysregulation and chronic inflammation. Inflammatory biomarkers such as interleukin-6 (IL-6) predict future cardiovascular risk among PWH (3,4). While people with HIV do have increased traditional risk factors for atherosclerosis such as dyslipidemia and smoking, identifying pathways that can mitigate the increased risk of HIV, even treated and well-controlled HIV, is critical as the population with HIV worldwide is aging and cardiovascular disease is of paramount significance clinically.

Recently, in the general population, clonal hematopoiesis of indeterminate potential (CHIP) has been associated with cardiovascular risk. CHIP is the expansion of a clonal population of hematopoietic stem cells with acquired somatic mutations at a variant allele fraction of >2% without hematologic cancer [National Cancer Institute Dictionary of genetics terms] (5). Murine models support the hypothesis that certain CHIP mutations (including in both TET2 and DNMT3a) are on the causal pathway to atherosclerosis via NLRP3 inflammasome activation of IL-1β and IL-6 (6-9). Human mendelian randomization studies also support this hypothesis, especially for TET2 mutations (10). Importantly, a human loss-of-function IL-6 receptor polymorphism, *IL6R* p.Asp358Ala, attenuates the risk of cardiovascular disease associated with large CHIP clones (11,12), suggesting that the mechanism by which CHIP increases cardiovascular risk is via IL-6 signaling associated with the NLRP3 inflammasome. Furthermore, a secondary analysis of the CANTOS trial, which compared the anti-IL-1β antibody canakinumab with placebo, suggested that those with TET2 mutations may response better to canakinumab (13). Even accounting for predicted cardiovascular risk from “traditional” risk factors and systemic inflammation with high-sensitivity C-reactive protein, CHIP is associated with increased cardiovascular mortality among individuals with carotid plaque.(14) While there are no CHIP-specific, evidence-based prevention strategies, a number of academic institutions have already established CHIP-Cardiovascular Disease Clinics to further address risk in this patient population. Furthermore, recent studies suggest that metformin, low-dose colchicine, and statins could be considered as therapeutic approaches to reduce or slow the risk of clonal expansion (15-20).

Prior research has demonstrated that HIV is associated with increased prevalence of CHIP, approximately double that of age-matched controls (21,22). As in the general population, CHIP has also been linked to IL-6 signaling among PWH (21). Among PWH, CHIP is detectable in blood samples obtained years prior to cardiovascular diagnosis and is associated with the subsequent development of cardiovascular disease (23). Whether there are HIV-specific processes that link increased risk of CHIP and increased risk of cardiovascular disease is unknown, but taken collectively, the shared common inflammatory pathways lend biologic plausibility to the hypothesis that these two processes may be tightly linked or synergistic among PWH.

Whether CHIP contributes to the development of subclinical atherosclerosis in PWH is unknown. We hypothesized that CHIP is associated with accelerated atherosclerosis among PWH via increased arterial inflammation and hematopoietic metabolic activity. To test this hypothesis, we measured carotid intima-media thickness to assess generalized atherosclerosis, soluble plasma biomarkers of inflammation, and ^18^F-FDG-PET activity among individuals with treated HIV.

## Methods

### Data Availability

Deidentified data are available upon reasonable request from the corresponding author.

### Design

This study is an observational study that includes both longitudinal and cross-sectional measurements that were performed as part of a longitudinal cohort study of people with HIV and baseline measures for clinical trials that enrolled people with HIV using the same imaging protocols.

### Participants

We included individuals ages 31-74 years old with diagnosed HIV, treated with antiretroviral therapy, and with undetectable viral load (<40 copies/ml).

### Primary Exposure Measurement

DNA was extracted from cryopreserved peripheral blood mononuclear cells stored in liquid nitrogen. CHIP was detected using a validated hybrid capture–based targeted gene panel (>1000x depth of coverage) that detects coding sequences for 22 common CHIP genes including: ASXL1, ASXL2, BRCC3, CBL, DNMT3A, ETNK1, GNAS, GNB1, IDH1, IDH2, JAK2, KIT, KRAS, MPL, NRAS, PPM1D, SETBP1, SF3B1, SRSF2, TET2, TP53, U2AF1(24). CHIP mutations were curated as previously described (25), and CHIP was defined as a variant allele fraction >2% (5). CHIP measurement was performed by a core laboratory blinded to all clinical variables and imaging data.

### Outcomes Measurements

To assess arterial inflammation and lymph node metabolic activity, we conducted ^18^F-FDG-PET at one time point according to our standard research protocol (26). ^18^F-FDG-PET reflects the degree of macrophage infiltration and foam cell formation, important drivers of atherosclerosis (27), as well as glycolytic pathways associated with atherosclerosis (28). Among PWH, lymph node metabolic activity can be a marker of HIV disease activity (29). The primary outcome was aortic inflammation as measured using the target-to-background ratio (TBR). Target-to-background ratio was calculated by dividing the standardized uptake value (SUV) in the aorta by the FDG uptake in the inferior vena cava, as previously described (30). Secondary outcomes included cervical and axiallary lymph node metabolic activity measured in terms of the mean standardized uptake value (SUV). FDG-PET images were transmitted securely to the reading center and interpreted by a core lab blinded to CHIP status and clinical variables.

As a substudy of the Study of the Consequences of the Protease-Inhibitor Era (SCOPE) cohort study, we conducted carotid ultrasound at two time points, baseline (time of CHIP measurement) and a second time point at a median 3.5 years later. (31). Ultrasounds were performed using high-resolution B-mode ultrasonography by trained research technicians according to our standard research protocol and then measured side-by-side. The primary outcome was mean of the 12-segement carotid IMT measured by an ultrasound technician blinded to CHIP status (which had not yet been measured) and other clinical variables consistent with the ARIC study and our prior work (31-33). We additionally assessed presence of carotid plaque, defined as a focal region of IMT greater than 1.5 mm, consistent with our prior work (31-33).

#### Correlative data

We measured inflammatory biomarkers that have previously been associated with both HIV and cardiovascular outcomes. Biomarkers (D-Dimer, fibrinogen IL-6, TNF, sCD14, sCD163, MCP1) were measured with a multiplex electrochemiluminescence assay. Markers of HIV reservoir from cell-associated DNA and RNA were previously measured in the subset with IMT (31).

Among the subset with FDG-PET CTs, we sent cryopreserved PBMCs to Accelevir Diagnostics. Accelevir Diagnostics performed HIV-1 Intact Proviral DNA Assay (IPDA®) to discriminate and separately quantify the frequencies of intact and defective persistent proviruses. The design and performance of assay have been described previously (34,35). Briefly, cryopreserved PBMCs were thawed and CD4+ T cells were isolated, with cell count, viability, and purity assessed by flow cytometry. RNA-free genomic DNA was then isolated from the recovered CD4+ T cells, with concentration and quality determined by fluorometry and ultraviolet-visible (UV/VIS) spectrophotometry, respectively. The IPDA® was performed, and data reported as proviral frequencies per million input CD4+ T cells. These procedures were performed by blinded operators using standard operating procedures. For the individuals with IMT, assessment of the viral reservoir was performed using cell-associated HIV RNA and DNA from enriched CD4+ T cells from cryopreserved PMBCS as previously described (31).

We also calculated the predicted 10-year atherosclerotic cardiovascular risk using the pooled cohort equations (36).

#### Statistical Approach

Non-parametric Wilcoxon Rank Sum tests were used for unadjusted comparisons of continuous variables between groups due to non-normal distributions of most variables of interest. Fisher’s exact test was used for comparison of categorical variables by group. For the primary cross-sectional outcomes, we used linear regression using complete-case analysis. We report two models based on uncertainty regarding which variables could contribute to development of CHIP: first a model adjusted for age and sex and second a model adjusted for age, sex, nadir CD4 count, smoking, treated hypertension, diabetes, LDL cholesterol, systolic blood pressure, and statin use. Given that nadir CD4 counts were not available for all participants and LDL could not be calculated for some individuals with high triglycerides, we repeated the expanded models, leaving out nadir CD4 count and LDL as sensitivity analyses. For longitudinal carotid intima-media thickness, we used linear mixed effects models with random effects per participant and accounting for differences in follow-up time. For carotid plaque, we used logistic regression models. Post-hoc secondary analyses stratifying by DNMT3A versus no DNMT3A CHIP were conducted for the primary outcomes. Statistical analyses were conducted using STATA Version 17.1. Statistical significance was defined as p<0.05 for the primary outcomes, and other outcomes were considered exploratory with confidence intervals and p-values reported without adjustment for multiple testing, given the shared primary hypothesis.

Institutional review committee approval was granted by the University of California San Francisco. All participants provided written informed consent.

## Results

We measured 437 samples from 230 PWH (median 2 samples per participant, interquartile range 1, 2). Participant characteristics at baseline are described in **Table 1**. We identified 32 individuals (13.9%) with CHIP mutations at a variant allele fraction >2% in at least one sample. Among those with CHIP, 30 individuals had one CHIP mutation and 2 had two mutations (in the same gene) present on both of their samples. The median variant allele fraction among those with CHIP was 2.8% (interquartile range 2.3%, 4.8%). Common mutations were in DNM3TA (n=21) and TET2 (n=6); less common mutations were in ASXL1 (n=2), PPM1D (n=2), BRCC3 (n=1), and TP53 (n=1). The two participants with two mutations each in the same gene had consistent results on both of their samples: two mutations in DNM3TA for one participant and two mutations in PPM1D for the other. One participant had CHIP based on a DNM3TA mutation on one sample (VAF 2.6%) and based on a TP53 mutation on a separate sample (VAF 2.7%).

**Table 1.**
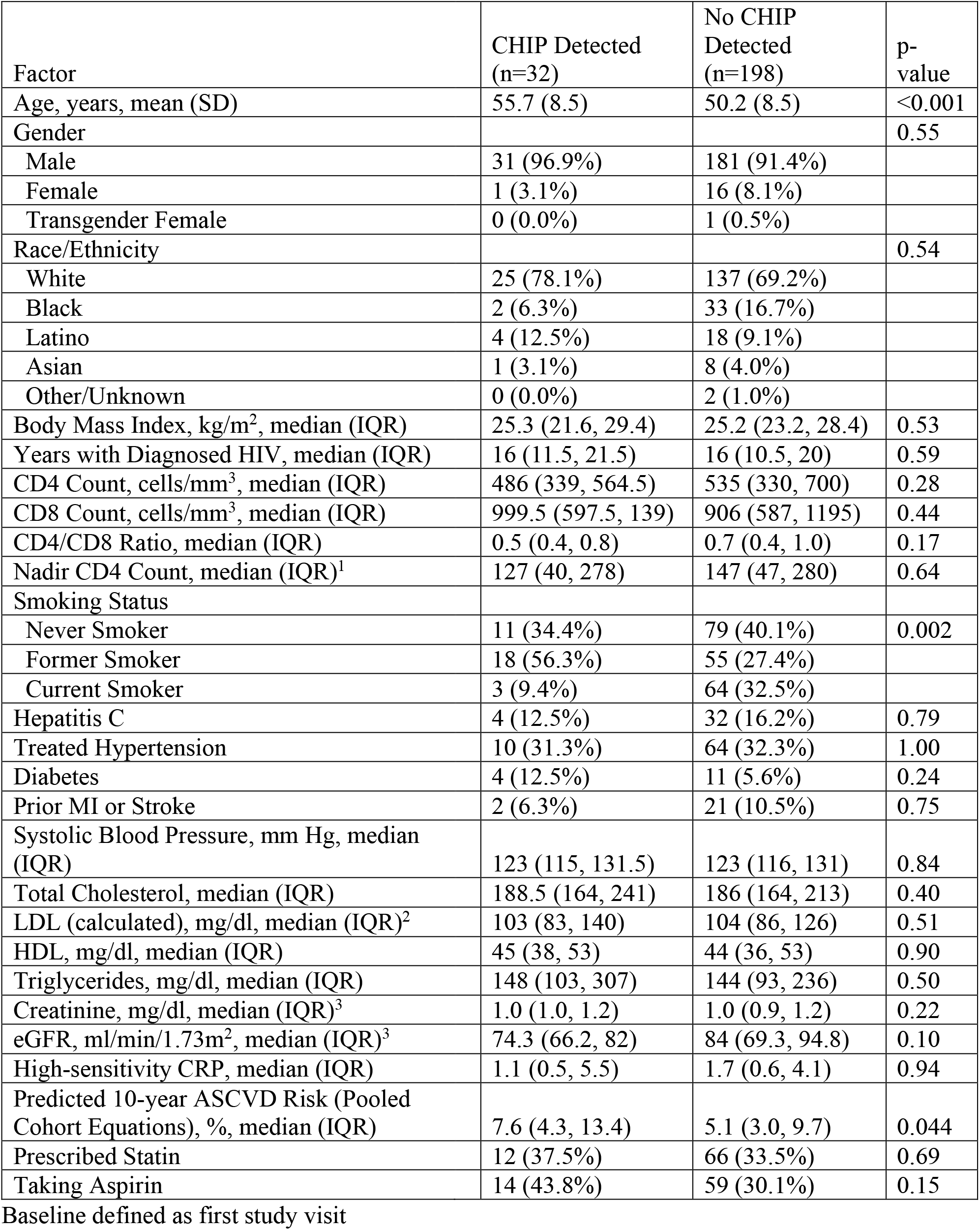
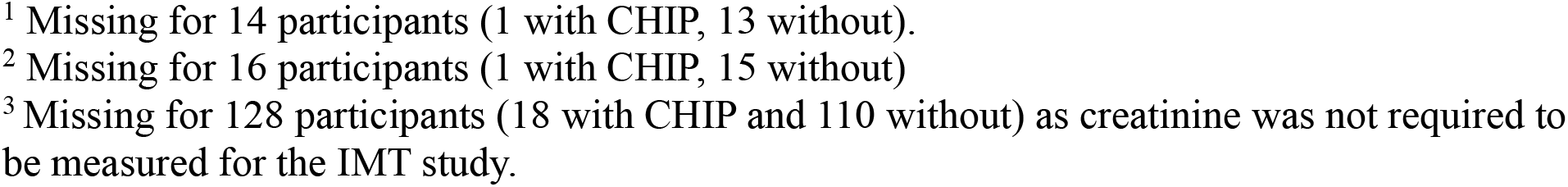
Participant Characteristics at Baseline by CHIP Status.

| Factor | CHIP Detected<br>(n=32) | No CHIP<br>Detected<br>(n=198) | p-value |
| --- | --- | --- | --- |
| Age, years, mean (SD) | 55.7 (8.5) | 50.2 (8.5) | <0.001 |
| Gender |  |  | 0.55 |
| Male | 31 (96.9%) | 181 (91.4%) |  |
| Female | 1 (3.1%) | 16 (8.1%) |  |
| Transgender Female | 0 (0.0%) | 1 (0.5%) |  |
| Race/Ethnicity |  |  | 0.54 |
| White | 25 (78.1%) | 137 (69.2%) |  |
| Black | 2 (6.3%) | 33 (16.7%) |  |
| Latino | 4 (12.5%) | 18 (9.1%) |  |
| Asian | 1 (3.1%) | 8 (4.0%) |  |
| Other/Unknown | 0 (0.0%) | 2 (1.0%) |  |
| Body Mass Index, kg/m<sup>2</sup>, median (IQR) | 25.3 (21.6, 29.4) | 25.2 (23.2, 28.4) | 0.53 |
| Years with Diagnosed HIV, median (IQR) | 16 (11.5, 21.5) | 16 (10.5, 20) | 0.59 |
| CD4 Count, cells/mm<sup>3</sup>, median (IQR) | 486 (339, 564.5) | 535 (330, 700) | 0.28 |
| CD8 Count, cells/mm<sup>3</sup>, median (IQR) | 999.5 (597.5, 139) | 906 (587, 1195) | 0.44 |
| CD4/CD8 Ratio, median (IQR) | 0.5 (0.4, 0.8) | 0.7 (0.4, 1.0) | 0.17 |
| Nadir CD4 Count, median (IQR)<sup>1</sup> | 127 (40, 278) | 147 (47, 280) | 0.64 |
| Smoking Status |  |  |  |
| Never Smoker | 11 (34.4%) | 79 (40.1%) | 0.002 |
| Former Smoker | 18 (56.3%) | 55 (27.4%) |  |
| Current Smoker | 3 (9.4%) | 64 (32.5%) |  |
| Hepatitis C | 4 (12.5%) | 32 (16.2%) | 0.79 |
| Treated Hypertension | 10 (31.3%) | 64 (32.3%) | 1.00 |
| Diabetes | 4 (12.5%) | 11 (5.6%) | 0.24 |
| Prior MI or Stroke | 2 (6.3%) | 21 (10.5%) | 0.75 |
| Systolic Blood Pressure, mm Hg, median (IQR) | 123 (115, 131.5) | 123 (116, 131) | 0.84 |
| Total Cholesterol, median (IQR) | 188.5 (164, 241) | 186 (164, 213) | 0.40 |
| LDL (calculated), mg/dl, median (IQR)<sup>2</sup> | 103 (83, 140) | 104 (86, 126) | 0.51 |
| HDL, mg/dl, median (IQR) | 45 (38, 53) | 44 (36, 53) | 0.90 |
| Triglycerides, mg/dl, median (IQR) | 148 (103, 307) | 144 (93, 236) | 0.50 |
| Creatinine, mg/dl, median (IQR)<sup>3</sup> | 1.0 (1.0, 1.2) | 1.0 (0.9, 1.2) | 0.22 |
| eGFR, ml/min/1.73m<sup>2</sup>, median (IQR)<sup>3</sup> | 74.3 (66.2, 82) | 84 (69.3, 94.8) | 0.10 |
| High-sensitivity CRP, median (IQR) | 1.1 (0.5, 5.5) | 1.7 (0.6, 4.1) | 0.94 |
| Predicted 10-year ASCVD Risk (Pooled Cohort Equations), %, median (IQR) | 7.6 (4.3, 13.4) | 5.1 (3.0, 9.7) | 0.044 |
| Prescribed Statin | 12 (37.5%) | 66 (33.5%) | 0.69 |
| Taking Aspirin | 14 (43.8%) | 59 (30.1%) | 0.15 |

Baseline defined as first study visit
<sup>1</sup> Missing for 14 participants (1 with CHIP, 13 without). <sup>2</sup> Missing for 16 participants (1 with CHIP, 15 without) <sup>3</sup> Missing for 128 participants (18 with CHIP and 110 without) as creatinine was not required to be measured for the IMT study.

As expected, older age was associated with ever detecting CHIP (OR 2.0 per decade older, 95% CI 1.3-3.01; p=0.002) and smoking status was also associated with CHIP (**Table 1**). Predicted 10-year atherosclerotic cardiovascular disease risk using the pooled cohort equations was higher among those with CHIP with a median of 8.7% (IQR 4.6, 13.4) compared to 5.1% (IQR 3.0, 10.0) among those without CHIP (p=0.044). No other clinical variables were associated with CHIP.

### Atherosclerosis Assessed Using Carotid Ultrasound

166 individuals (n=23 with CHIP) completed a baseline carotid ultrasound and 161 (21 with CHIP) completed a follow-up carotid ultrasound (**Supplemental Table 1**). At baseline, CHIP was not associated with carotid intima-media thickness (mean IMT 1.093 mm with CHIP compared to 1.001 mm without CHIP; difference 0.092 mm; 95% CI −0.054 to 0.239 mm; p=0.21; **Figure 1**). Accounting for age and sex did not meaningfully change the results (−0.020 mm; 95% CI −0.15 to 0.11 mm; p=0.76), nor did the model adjusted for age, sex, nadir CD4 count, smoking, treated hypertension, diabetes, LDL cholesterol, systolic blood pressure, and statin use (0.023 mm; 95% CI −0.13 to 0.18; p=0.77). Sensitivity analyses using only CHIP detected concurrently with IMT measurement had similar results (unadjusted p=0.17).

**Figure 1.**
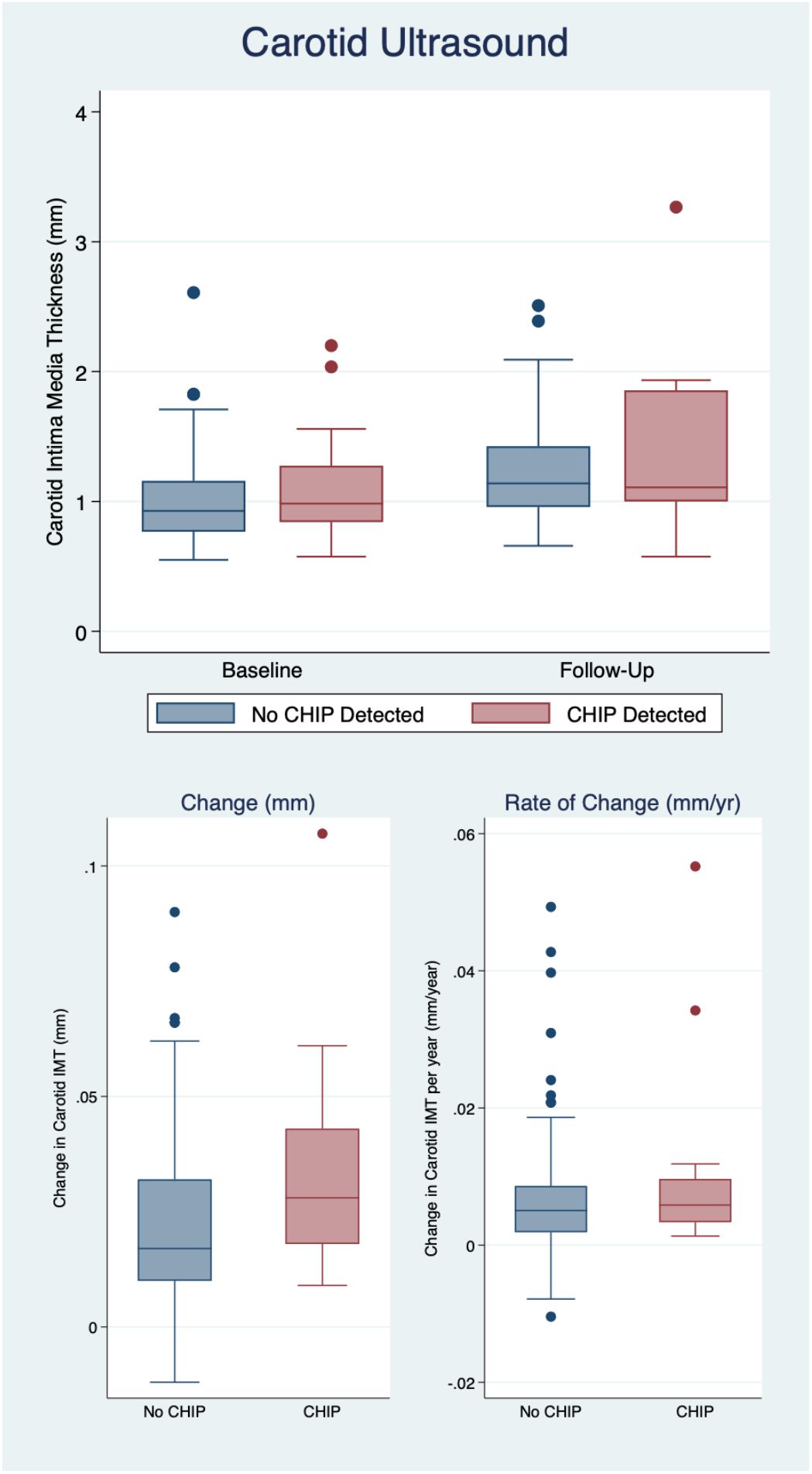
Longitudinal Carotid Intima Media Thickness by CHIP. Figure Legend: Boxplots of carotid IMT at baseline and follow-up, a median 3.5 years after baseline, (top) and change in IMT (bottom panel) by CHIP status at Baseline and Follow-Up. Those with CHIP are older and had higher predicted cardiovascular risk by the pooled cohort equations, which likely explains the greater IMT among those with CHIP at both baseline and follow-up; there was no difference in the change in IMT by CHIP status (bottom panel).

In a post-hoc analysis, DNMT3A CHIP was associated with slightly greater IMT thickness at baseline than non-CHIP (0.032 mm greater, 95% CI 0.009 to 0.055; p=0.007), but non-DNMT3A CHIP was not (−0.002 mm greater, 95% CI −0.020 to 0.015; p=0.79). However, accounting for age and sex attenuated the difference between DNTM3A CHIP and non-CHIP (0.009 mm greater, 95% CI −0.012 to 0.031; p=0.39) with similar results adjusted for additional potential confounders (age, sex, nadir CD4 count, smoking, treated hypertension, diabetes, LDL cholesterol, systolic blood pressure, and statin use; 0.012 mm greater, 95% CI −0.011 to 0.035; p=0.32).

Over a median follow up of 3.5 years, IMT thickness increased a mean 0.23 mm. Mean IMT did not increase more among those with CHIP (0.028 mm greater with CHIP, 95%CI −0.17 to 0.22 mm; p=0.78; **Figure 1**) or faster (0.0068 mm/year; 95% CI −0.037 to 0.051 mm/year; p=0.76). We did not detect evidence of faster atherosclerosis in models accounting for age and sex (−0.033 mm/year more with CHIP; 95% CI −0.11 to 0.039 mm/year; p=0.37) or with an expanded list of potential confounders as above (−0.026 mm/year; 95% CI −0.087 to 0.035; p=0.41).

In post-hoc analysis, DNMT3A CHIP was associated with greater increase in IMT (0.025 mm greater increase than non-CHIP; 95% CI 0.008 to 0.04; p=0.004), and non-DNMT3A CHIP was associated with smaller increase than non CHIP (−0.017 mm than non CHIP, 95% CI −0.32 to −0.003; p=0.022). DNMT3A CHIP was associated with faster IMT expansion compared to non-CHIP (0.057 mm/year faster; 95% CI 0.002 to 0.009; p=0.003) and non-DNTMA CHIP was associated with slightly slower IMT expansion compared to non CHIP (−0.003 mm/year; 95% CI −0.07 to 0.00; p=0.056). These results were consistent with adjustment for age and sex and accounting for additional potential confounders (age, sex, nadir CD4 count, smoking, treated hypertension, diabetes, LDL cholesterol, systolic blood pressure, and statin use).

Similarly, CHIP was not strongly associated with carotid plaque at baseline (15/23, 65% vs 77/143, 54%; OR 1.61, 95% CI 0.64 to 4.02; p=0.31). We did not find evidence for an association between CHIP and carotid plaque in models adjusted for age and sex (OR 0.90 for prevalent plaque at baseline, 95% CI 0.32-2.53) or more fully adjusted (OR 0.99 95% CI 0.29-3.36). CHIP was not strongly associated with development of incident plaque among those without plaque at baseline (3/7 (43%) with CHIP vs 25/66 (38%) without CHIP; OR=1.25, 95% CI 0.26 to 6.07; p=0.78). Results were similar with the adjusted models (OR 0.91 adjusted for age and sex, 95%CI 0.17-4.76; p=0.91; and OR 1.60 for full model, 95% CI 0.25-10; p=0.62), and in sensitivity analyses only using detectable CHIP at baseline.

### FDG-PET CT Findings

There were 80 individuals (12 with CHIP) with adequate quality FDG-PET CT images to calculate the aortic target-to-background ratio (TBR) (**Supplemental Table 2**). CHIP was not associated with aortic inflammation (mean TBR±standard deviation: 3.02±0.9 with CHIP vs 3.06±0.9 without CHIP; difference −0.039, 95% CI −0.58 to 0.50; p=0.89; **Figure 2**). This was unchanged in the model that included age and sex (difference 0.094, 95% CI −0.048 to 0.67; p=0.90) or the model that included additional potential confounders (difference 0.36, 95% CI −0.23 to 0.95; p=0.23). In a post-hoc analysis subsetting DNMT3A and non-DNMT3A CHIP, we did not find differences in aortic TBR (DNMT3A vs non CHIP −0.026, 95% CI −0.64 to 0.58; p=0.93 and non-DNTM3A CHIP vs non CHIP −0.068, 95% CI −1.08 to 0.95; p=0.89).

**Figure 2.**
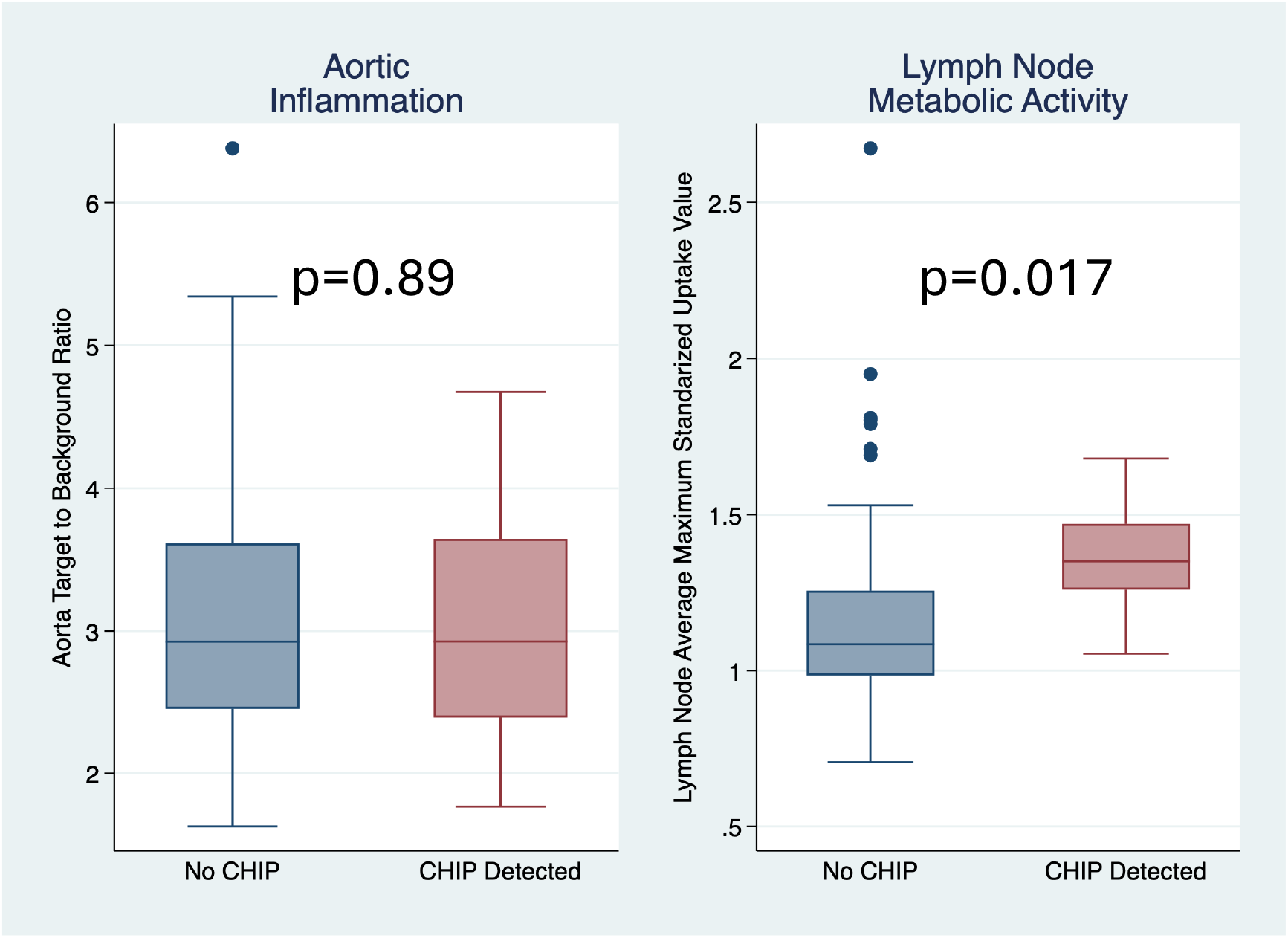
Aortic Inflammation and Lymph Node Metabolic Activity. Figure Legend: Aortic inflammation (n=79, 12 with CHIP) was not increased among those with CHIP detected (p=0.89); in contrast, lymph node metabolic activity (n=50, 9 with CHIP) was increased (p=0.017).

Among those whose lymph nodes were assessed (n=50, CHIP=9), CHIP was associated with increased lymph node metabolic activity (SUVmax 1.37±0.19 with CHIP vs 1.20±0.40 without CHIP; p=0.017). The difference in lymph node activity was similar for both DNMT3a and non-DNT3A CHIP, 0.23 and 0.26, respectively. This association was no longer statistically significant with adjustment for age, sex, and potential clinical confounders or when assessed using the target-to-background ratio in reference to blood pool activity.

### CHIP Is Not Associated with Inflammatory Biomarkers

We had previously measured inflammatory biomarkers in 150 participants who had carotid IMT and were included in this study (CHIP=21). We did not find that CHIP was associated with any of the inflammatory biomarkers we assessed (**Table 2**).

**Table 2.**
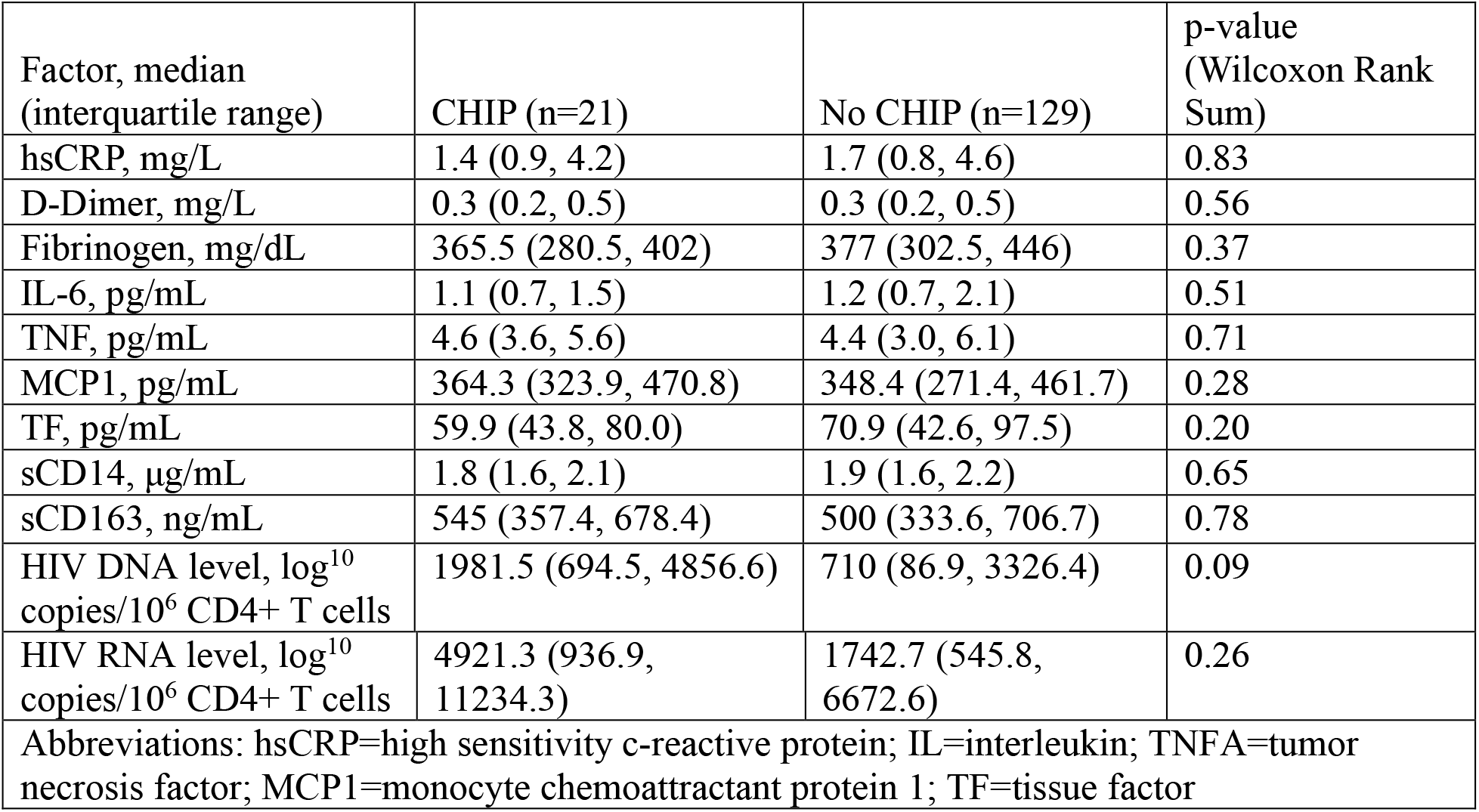
Biomarkers of Inflammation by CHIP status.

| Factor, median<br>(interquartile range) | CHIP (n=21) | No CHIP (n=129) | p-value<br>(Wilcoxon Rank<br>Sum) |
| --- | --- | --- | --- |
| hsCRP, mg/L | 1.4 (0.9, 4.2) | 1.7 (0.8, 4.6) | 0.83 |
| D-Dimer, mg/L | 0.3 (0.2, 0.5) | 0.3 (0.2, 0.5) | 0.56 |
| Fibrinogen, mg/dL | 365.5 (280.5, 402) | 377 (302.5, 446) | 0.37 |
| IL-6, pg/mL | 1.1 (0.7, 1.5) | 1.2 (0.7, 2.1) | 0.51 |
| TNF, pg/mL | 4.6 (3.6, 5.6) | 4.4 (3.0, 6.1) | 0.71 |
| MCP1, pg/mL | 364.3 (323.9, 470.8) | 348.4 (271.4, 461.7) | 0.28 |
| TF, pg/mL | 59.9 (43.8, 80.0) | 70.9 (42.6, 97.5) | 0.20 |
| sCD14, µg/mL | 1.8 (1.6, 2.1) | 1.9 (1.6, 2.2) | 0.65 |
| sCD163, ng/mL | 545 (357.4, 678.4) | 500 (333.6, 706.7) | 0.78 |
| HIV DNA level, log<sup>10</sup><br>copies/10<sup>6</sup> CD4+ T cells | 1981.5 (694.5, 4856.6) | 710 (86.9, 3326.4) | 0.09 |
| HIV RNA level, log<sup>10</sup><br>copies/10<sup>6</sup> CD4+ T cells | 4921.3 (936.9,<br>11234.3) | 1742.7 (545.8,<br>6672.6) | 0.26 |
| Abbreviations: hsCRP=high sensitivity c-reactive protein; IL=interleukin; TNFA=tumor<br>necrosis factor; MCP1=monocyte chemoattractant protein 1; TF=tissue factor |  |  |  |

### Viral Reservoir

We investigated whether CHIP was associated with markers of the viral reservoir using two approaches. Among 150 individuals who had completed longitudinal carotid IMT (21 with CHIP), CHIP was not strongly associated with cell-associated HIV DNA (unadjusted p=0.08) or with HIV RNA (unadjusted p=0.25; **Table 2**). Among 78 with FDG PET CT data (12 with CHIP), CHIP is not associated with the intact proviral reservoir assessed by IPDA (p=0.72).

## Discussion

In this study, we tested the hypotheses that CHIP increases cardiovascular risk among PWH by impact on: 1) atherosclerosis burden and 2) arterial inflammation. We used validated imaging measures to assess carotid intima media thickness and plaque (with carotid ultrasound), and arterial inflammation (using ^18^F FDG-PET CT). Notably, we did not find evidence for an association between CHIP and subclinical markers of atherosclerosis or atherosclerosis progression among people with HIV at elevated cardiovascular risk (**Visual Abstract**), although we did find potential differences by CHIP subtype (DNMT3A versus non-DNMT3A CHIP) on carotid IMT. Thus, our key findings directly contradicted our study hypotheses: we expected to find greater IMT thickness, faster IMT progression, and higher levels of arterial inflammation among those with CHIP, yet our overall study findings were negative (except for post-hoc analysis of DNMT3A CHIP and carotid atherosclerosis).

Our study hypotheses were based on several observations made in the general population of individuals living without HIV. In a study that included four case-control studies with a total of 8,255 participants, CHIP detected in peripheral blood cells was associated with increased risk of myocardial infarction (6). In the Progression of Early Subclinical Atherosclerosis study, CHIP was not associated with sublinical atherosclerosis burden across multiple vascular territories (carotid, femoral, and coronary) after adjustment for age, sex, and conventional risk factors, but CHIP was associated with incident femoral atherosclerosis at 3 years, with a dose-dependent response to clone size (37). Interestingly, that study was not able to detect differences in incident carotid atherosclerosis or carotid plaque progression. A recent study called into question the causal direction of the association between clonal hematopoeisis and atherosclerosis by demonstrating increased hematopoietic stem cell division rates in both mice and humans (38); thus CHIP may result from atherosclerosis rather than promote atherosclerosis.

In contrast to studies in the general population, studies looking at subclinical atherosclerosis in PWH and CHIP have not demonstrated consistent findings. In a subset of 188 participants from the Multicenter AIDS Cohort study, using a VAF>1%, coronary stenosis greater than 50% was more common in PWH and CHIP than those without CHIP (39). In contrast, in the Copenhagen Comorbidity in HIV Infection (COCOMO) study, CHIP defined as VAF>2% was common (49/190 individuals, 26%) but was not associated with presence of CAD or obstructive CAD (40). In a study of 4490 PWH at low-to-intermediate cardiovascular risk in the Randomized Trial to Prevent Vascular Events in HIV (REPRIEVE) trial, CHIP was not associated with MACE, but there was a dose-dependent response with large clones (VAF≥10%) associated with increased risk (41).

There are several plausible explanations for the contradictory findings across these different studies. Differences in the relative prevalence of specific mutations within CHIP genes (R882 mutations in DNMT3, for example), may impact the findings (42). Secondly, HIV may be a different context – the baseline level of chronic inflammation in the setting of HIV may make the relative contribution of CHIP less relevant to atherogenesis specifically among PWH. The inflammatory environment of HIV may also contribute to the development of CHIP, raising questions about the directionality of association. Several studies suggest CHIP may be a marker of more severe HIV disease (41), which may complicate associations with subclinical atherosclerosis. Finally, the modest sample sizes among most studies of PWH may contribute to the heterogeneity of findings.

One possible explanation underlying our findings in this study is that the most common mutation in our cohort was in DNMT3, whereas TET2 mutations (which were less common in our cohort) have the strongest evidence for atherosclerosis in humans. DNMT3A, DNA methyltransferase 3 alpha, is an epigenetic regulator; inactivation of DNMT3 in macrophages may increase cardiac fibrosis via interactions with cardiac fibroblasts, ultimately leading to heart failure (43,44). Although some murine models have linked the downstream inflammation from both DNMT3A and TET2 (9), the overlap of downstream effects in humans is less certain (45). In the UK Biobank, only non-DNMT3A CHIP in the general population is associated with incident coronary artery disease (12). However, our results suggest the opposite—DNMT3A CHIP was associated with greater IMT at baseline and faster IMT progression, while non-DNMT3A (which included TET2 and others) was not.

We also did not find evidence that CHIP is associated with higher levels of the inflammatory markers that we studied, including those previously associated with atherosclerosis among PWH. It was surprising that IL-6 levels were not higher among PWH with CHIP compared to those without CHIP because this has been previously observed in the general population in CANTOS (13), and in two prior studies that included PWH (21,46). Studies within HIV have been more heterogenous, with both the MACS and COCOMO studies not demonstrating associations between CHIP and inflammatory markers (39,40). In contrast, among individuals with a history of advanced HIV and a history of immune reconstitution inflammatory syndrome (IRIS), PWH had a prevalence of CHIP of 27% and the history of IRIS was associated with CH along with higher inflammatory markers (46). It could be that immune dysregulation and chronic inflammation in the setting of treated and suppressed HIV blunts or washes out the effect of NLRP3 inflammasome activation specifically from CHIP in this population.

One hypothesis-generating finding is increased lymph node metabolic activity among PWH with CHIP, which was not one of our primary hypotheses. Lymphadenopathy is a common finding among people with HIV, and metabolic activity in lymph nodes assessed by ^18^F-FDG-PET is associated with higher levels of HIV disease activity and worse immunosuppression (29). Maintenance of the HIV reservoir in lymph nodes is due to clonal proliferation of cells infected with HIV before initiation of antiretroviral therapy, with evidence of trafficking of cells between peripheral blood and hematopoietic compartments, rather than ongoing HIV replication in lymph nodes (47). Whether the increased metabolic activity we observed reflects recruitment of macrophages to lymph node harbors of CHIP-mutated and/or HIV-infected cells or reflects the localized HIV reservoir would require additional studies that include lymph node biopsies of PWH. While not statistically significant, there was a higher level of HIV DNA copies per CD4+ T cell among those with CHIP. This finding of increased lymph node metabolic activity (possibly in association with greater reservoir activity) also raises the possibility that CHIP in HIV could be a precursor of eventual lymphoma, as has been reported after chimeric antigen receptor T cell therapy selecting for CHIP mutations (48). With this cross-sectional analysis, the temporal direction of this potential association is unclear.

### Limitations

This mechanism-oriented study has several major limitations. There were notable differences between those with and without CHIP, such as older age, that may have made it difficult to disentangle the effects of CHIP from other factors. As the study was conducted in San Francisco where approximately 95% of people with HIV are men, the study only included 7% women. Because this study leveraged measurements obtained for other study protocols, we did not have carotid ultrasounds, FDG PET/CTs, or biomarkers on all study participants in whom we had measured CHIP. In addition, the follow-up time may have been too short to detect acceleration of atherosclerosis by CHIP status. Our study was also relatively small to consider the differential impact that different CHIP mutations may have on atherosclerosis and inflammation, but we did conduct post-hoc analyses comparing DNMT3A and non-DNMT3A CHIP; combining all CHIP mutations may have made it harder to detect effects that are mutation specific, especially since we had so few individuals with TET2 mutations. We did not use *in vivo* assays to assess cardiac fibrosis such as cardiac magnetic resonance imaging with parametric mapping, which may have identified DNTM3A-associated cardiac fibrosis.

### Conclusions

Within the SCOPE cohort of PWH, CHIP mutations were not associated with subclinical atherosclerosis, arterial inflammation, or inflammatory biomarkers. The link between lymph node metabolic activity, CHIP, and its potential relationship with the HIV reservoir merits validation in a follow-up study. Further investigation into the role of CHIP in the development of cardiovascular disease among PWH would benefit from larger sample sizes and longer followup, perhaps including clinical events.

## Data Availability

Deidentified data are available upon reasonable request from the corresponding author.

